# Optimising COVID-19 Episode Identification Using Serology and PCR/Rapid Antigen Testing: Insights from the BRACE Trial

**DOI:** 10.1101/2025.03.12.25323795

**Authors:** Ellie McDonald, Laure F Pittet, Marc Bonten, Anthony Byrne, John Campbell, Julio Croda, Margareth Pretti Dalcolmo, Andrew Davidson, Glauce dos Santos, Kaya Gardiner, Amanda Gwee, Bruno Araújo Jardim, Marcus Vinícius Guimaraes Lacerda, Michaela Lucas, David J Lynn, Laurens Manning, Helen S Marshall, Kirsten P Perrett, Cristina Prat-Aymerich, Marco AM Puga, Peter C Richmond, Jesús Rodríguez-Baño, Ushma Wadia, Adilia Warris, Nicholas J Wood, Nigel Curtis, Nicole L Messina, the BRACE Trial Consortium Group

## Abstract

**Background:** Accurately identifying COVID-19 episodes was crucial during the pandemic for evaluating interventions. Results from diagnostic tools like PCR, rapid antigen test (RAT) and serology are affected by factors such as timing of tests and vaccination status. The BRACE trial developed an algorithm integrating these diagnostic tools for illness episode classification.

**Methods:** In the BRACE trial, 3988 participants reported 5512 febrile/respiratory illness episodes and provided longitudinal blood samples over one year. SARS-CoV-2 diagnosis relied on a three-component algorithm: (1) a serology algorithm assessing anti-SARS-CoV-2 nucleocapsid antibody seroconversion, (2) a PCR/RAT algorithm, and (3) an episode interpretation algorithm combining serology and PCR/RAT results to categorise episodes as COVID-19, Not COVID-19 or Uncertain. The algorithms accounted for vaccination status and timing of testing relative to symptom onset to refine episode classifications.

**Results:** Of 5512 illness episodes, 890 (16%) were classified as COVID-19, 3852 (70%) as Not COVID-19, and 770 (14%) as Uncertain. Compared to relying solely on PCR/RAT results, integrating serology in the algorithm reduced the proportion of Uncertain classifications by more than half. Among the COVID-19 episodes, 89% were identified by positive PCR/RAT results, and the remaining 11% (with missing or negative PCR/RAT tests) were identified by serology. Discordance between PCR/RAT and serology occurred in 13% of episodes.

**Conclusion:** An algorithm integrating PCR/RAT and serology results in the context of test timing and vaccine status enabled the accurate identification of COVID-19 episodes and minimised the number of episodes that would otherwise have been classified as Uncertain.

## Background

Accurately identifying COVID-19 episodes was vital for evaluating interventions in the pandemic. The performance of diagnostic tests for SARS-CoV-2, such as respiratory swab testing and serology, is influenced by the timing of testing relative to symptom onset, as well as prior COVID-19 vaccinations.

In the BRACE trial, comprehensive symptom data were collected alongside SARS-CoV-2 testing.[1–3] Here, we describe the algorithm developed during the BRACE trial, intended to more precisely identify COVID-19 episodes using illness episode data and diagnostic test results.

To account for test timing in relation to symptom onset and any effect of COVID-19 vaccination, a three-component interpretation algorithm was developed. This incorporated serological result interpretation, PCR/rapid antigen test (RAT) respiratory swab test interpretation, and finally episode classification to COVID-19, Not COVID-19 or Uncertain categories.

In addition to evaluating the sensitivity and specificity of PCR/RAT testing compared to serology for SARS-CoV-2, and the frequency of discordant results, we assessed the impact of each component of the algorithm on episode classification.

## Methods

### Participants and study design

The BRACE trial (NCT04327206) was a phase 3 international randomised controlled trial to assess the impact of BCG vaccination on the prevalence of COVID-19 among healthcare workers.[1–3] Trial outcomes relied on accurate ascertainment of the onset of a participant’s first episode of COVID-19.

The trial was designed early in the pandemic, with recruitment starting in March 2020. This study included the 3988 participants recruited to the BRACE trial from May 2020 to April 2021 from Australia, the Netherlands, Spain, the UK and Brazil. Selection of COVID-19 symptoms to be collected and diagnostic tests for SARS-CoV-2 were informed by best practice and government policies at the time.

### Illness episode ascertainment

Recent illness symptoms were ascertained weekly via a custom-built smartphone app or direct communication (phone call or text). During illness episodes, symptoms were recorded daily, and participants were prompted to undergo SARS-CoV-2 respiratory swab testing using PCR (via government testing centres) or RAT (self-administered). Additionally, more comprehensive questionnaires were administered at the start of the trial and quarterly throughout follow-up. These collected information about COVID-19-specific vaccinations (date and vaccine type). Blood samples were collected at baseline and 3, 6, 9 and 12 months post-randomisation to measure anti-SARS-CoV-2 nucleocapsid (NCP) antibodies using the Roche Cobas Elecsys anti-SARS-CoV-2 assay [4, 5] at the Victorian Infectious Diseases Reference Laboratory. For seroconversion, a cut-off index (COI) of ≥1.0 was used as per manufacturer’s instructions.[6, 7]

### Serological interpretation algorithm

The serology algorithm produced one of four possible outcomes: Seroconversion, No seroconversion, Indeterminate or No result (where a blood sample was unavailable). For each illness episode, the most recent blood sample collected at least seven days prior to symptom onset (pre-episode sample), and the first and second blood samples collected after symptom onset were identified. The NCP serology result from these samples were used in the serological interpretation algorithm. The algorithm considered the timing of blood sample in relation to the illness episode, any prior administration of the *CoronaVac* (Sinovac Biotech) COVID-19 vaccine (a whole inactivated virus vaccine which can induce anti-NCP antibodies) and change in anti-NCP antibody titre (in cases of positive-to-positive serology results). A detailed description of the serological algorithm is provided in Supplementary Figure 1.

### PCR/RAT interpretation algorithm

The PCR/RAT interpretation algorithm yielded three possible outcomes: Positive, Negative, or No result. Both PCR and RAT positive results were considered confirmation of SARS-CoV-2 infection if the timing of the test fell within defined symptomatic illness parameters. Positive PCR results were considered valid if the sample was tested within three days prior to 10 days (RAT) or 21 days (PCR) after the start, or within seven days from the end, of an illness episode. A negative PCR was considered ‘Negative’, but a negative RAT was classified as ‘No result’ due to the lower sensitivity of RAT testing.[8] A detailed description of the PCR/RAT interpretation algorithm is provided in Supplementary Figure 2.

### Episode interpretation algorithm

The episode interpretation algorithm produced three possible classifications for each illness episode: COVID-19, Not COVID-19 or Uncertain (which included episodes for which results were missing, or indeterminate). The results of the serological and PCR/RAT algorithms were combined to categorise illness episodes using the episode interpretation algorithm, detailed in Figure 1. Additional conditions were then applied which evaluated other COVID-19 tests and episode results within the same timeframe.

**Figure 1.**
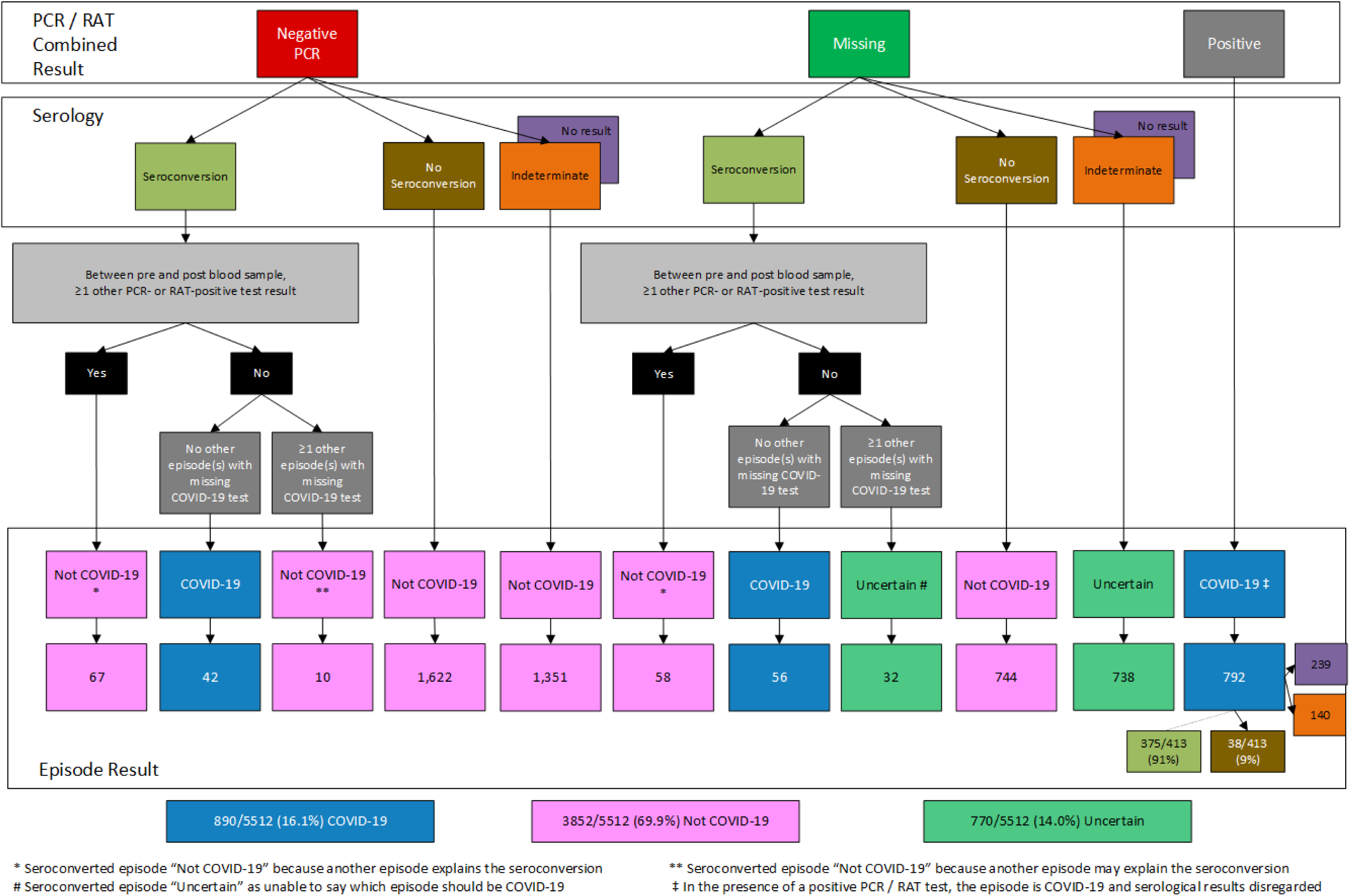
Episode interpretation algorithm: the detail

## Results

Of the 3988 participants, 2559 reported at least one episode of illness in the year post-randomisation, with a total of 5512 episodes.

Of the 5512 episodes, 890 (16%) were classified as COVID-19, 3852 (70%) as Not COVID-19 and 770 (14%) as Uncertain (Figure 1). The participant flow from the combined PCR/RAT result and from the pre-episode NCP serology to the final episode classification are shown in Figure 2 and Supplementary Figure 3 respectively.

**Figure 2.**
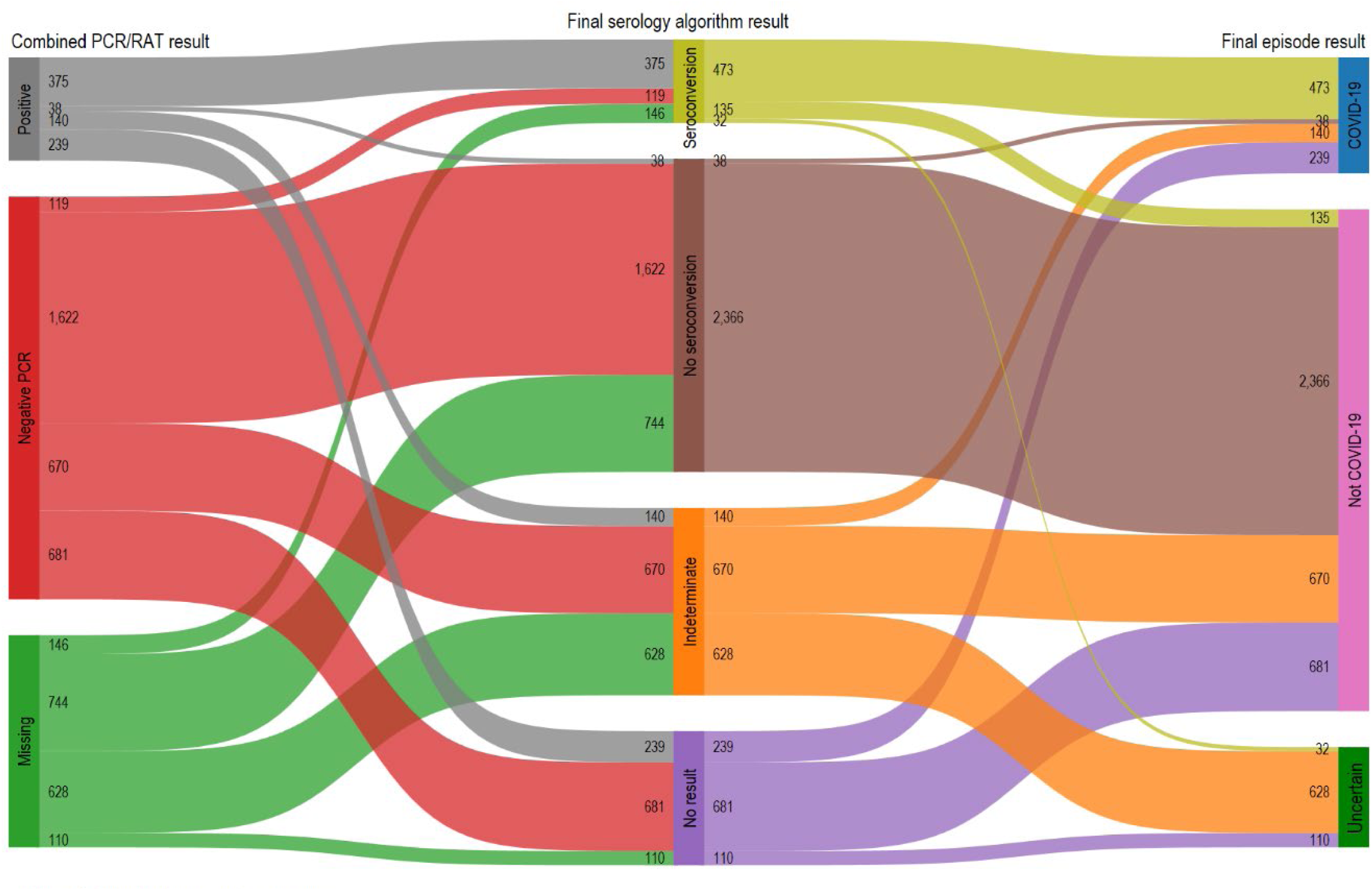
Episode interpretation outcome

Of the 890 episodes defined as COVID-19, 792 (89%) had a positive PCR or RAT test result. Serology results were concordant (seroconversion) in 375 (47.4%) episodes, discordant (no seroconversion) in 38 (4.8%), and Indeterminate or No result in 379 (47.9%) (Figure 2). The remaining 98 COVID-19 episodes comprised 56 (6.3%) with a missing PCR/RAT result and 42 (4.7%) with a negative PCR result (Supplementary Figure 4). The discordant PCR/RAT and serology results were not attributable to post-episode NCP titres being just above the threshold for seroconversion (Figure 3). The post- episode NCP titres in relation to the time interval between the episode start and post-episode blood collection for the seroconverted PCR negative and PCR/RAT positive episodes are shown in Supplementary Figure 5. Median post-episode NCP titres were higher in seroconverting episodes with negative PCR results compared to those with positive PCR/RAT results (median 62.2, IQR: 15.9– 122.9 vs. 41.3, IQR: 14.3–101.4; p=0.3). Relevant to this, median time between episode onset and post-episode serological testing was higher in the negative PCR episode group compared to the positive PCR/RAT group (median 76.5 days, IQR: 33-102 days vs. 61 days, IQR: 37-82 days; p=0.1).

**Figure 3.**
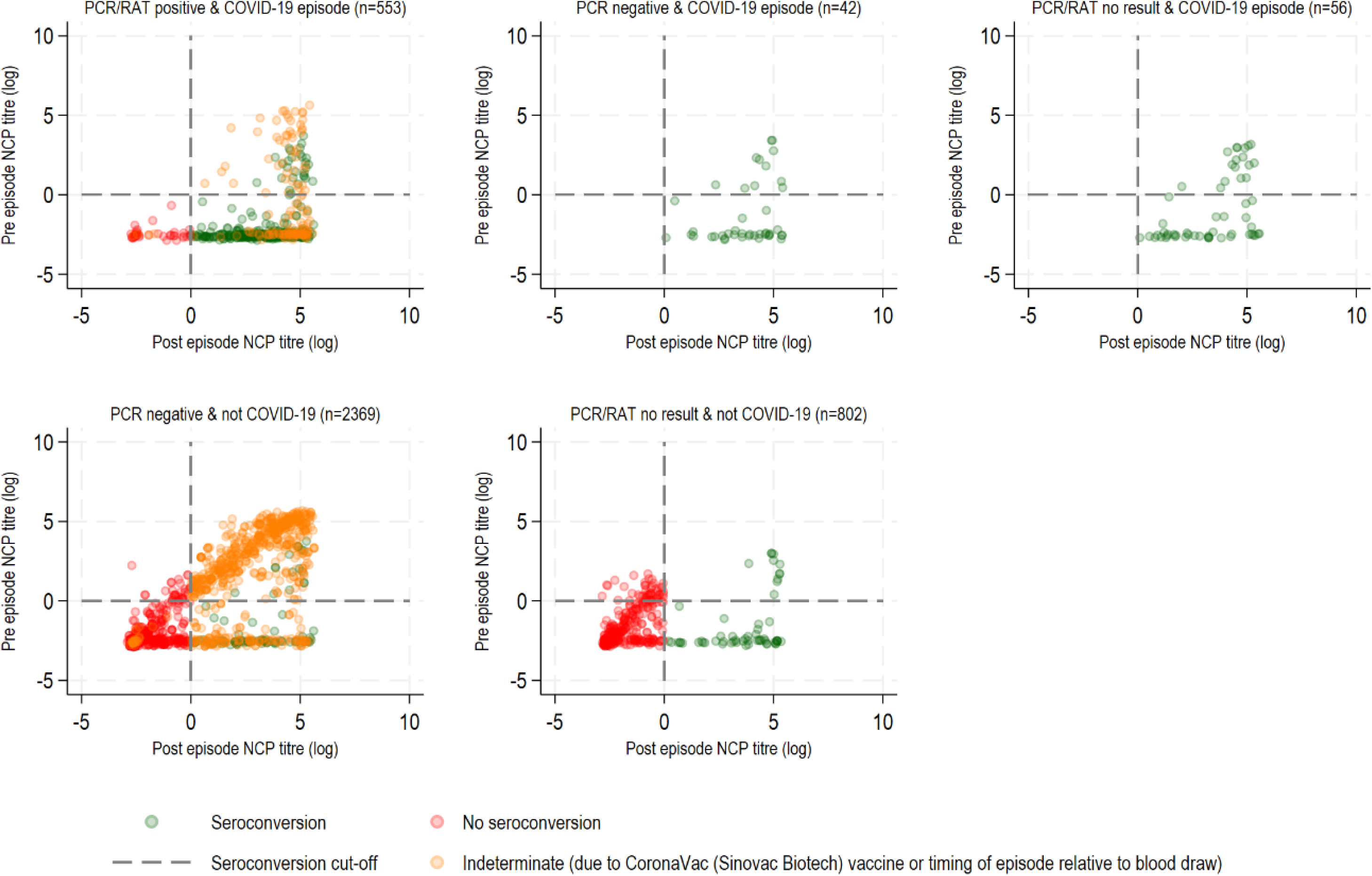
Pre and post NCP titre serology results by PCR/RAT and episode results

There were 305 episodes that had a negative RAT test. Of the RAT-negative episodes without a PCR result, 11/179 (6.2%) were categorised as COVID-19 based on seroconversion (Supplementary Figure 6). In comparison, of the episodes without a PCR result, 56/858 (7.0%) were categorised as COVID-19 based on seroconversion (Figure 3).

Among the 3852 episodes classified as Not COVID-19 by the final episode algorithm, 3050 (77%) had a negative PCR result (Figure 2 & Supplementary Figure 3). Of these, 1622 (53.2%) had concordant negative serology and PCR results (Figure 2). Additionally, 1351 (44.3%) episodes had a missing NCP serology result, but a negative PCR result. The remaining 77 (2.5%) episodes were discordant (seroconversion with a negative PCR result). However, the positive NCP serology result in these cases was attributed to participants’ other PCR- or RAT-positive episodes within the same timeframe. For the remaining 802 (20%) Not COVID-19 episodes with a missing PCR result, 744 had no seroconversion between pre- and post-episode blood samples. These episodes would have been classified as Uncertain if the trial had relied on PCR/RAT results only. In the remaining 58 (7.2%) episodes, participants seroconverted, but the episode algorithm attributed this to another PCR- or RAT-positive episode.

Of the 770 episodes classified as Uncertain in the final episode result, 738 (95.8%) had both missing or indeterminant serology and PCR/RAT results. Of the remaining 32 episodes, participants seroconverted but the episodes were categorised as Uncertain because the participants had multiple episodes with missing PCR/RAT results within the same seroconversion timeframe. This precluded the attribution of the COVID-19 diagnosis to one of the episodes.

At the participant level, Uncertain result episodes were similarly distributed between every combination of COVID-19 and Not COVID-19 episodes (Figure 4). The sequential distribution of COVID-19, Not COVID-19 and Uncertain result episodes for each participant demonstrates the progression from algorithm processing to the final trial outcomes. It highlights the points at which participants were censored, which occurred at one of two events: their first COVID-19 episode or their first episode with an Uncertain result (Figure 5).

**Figure 4.**
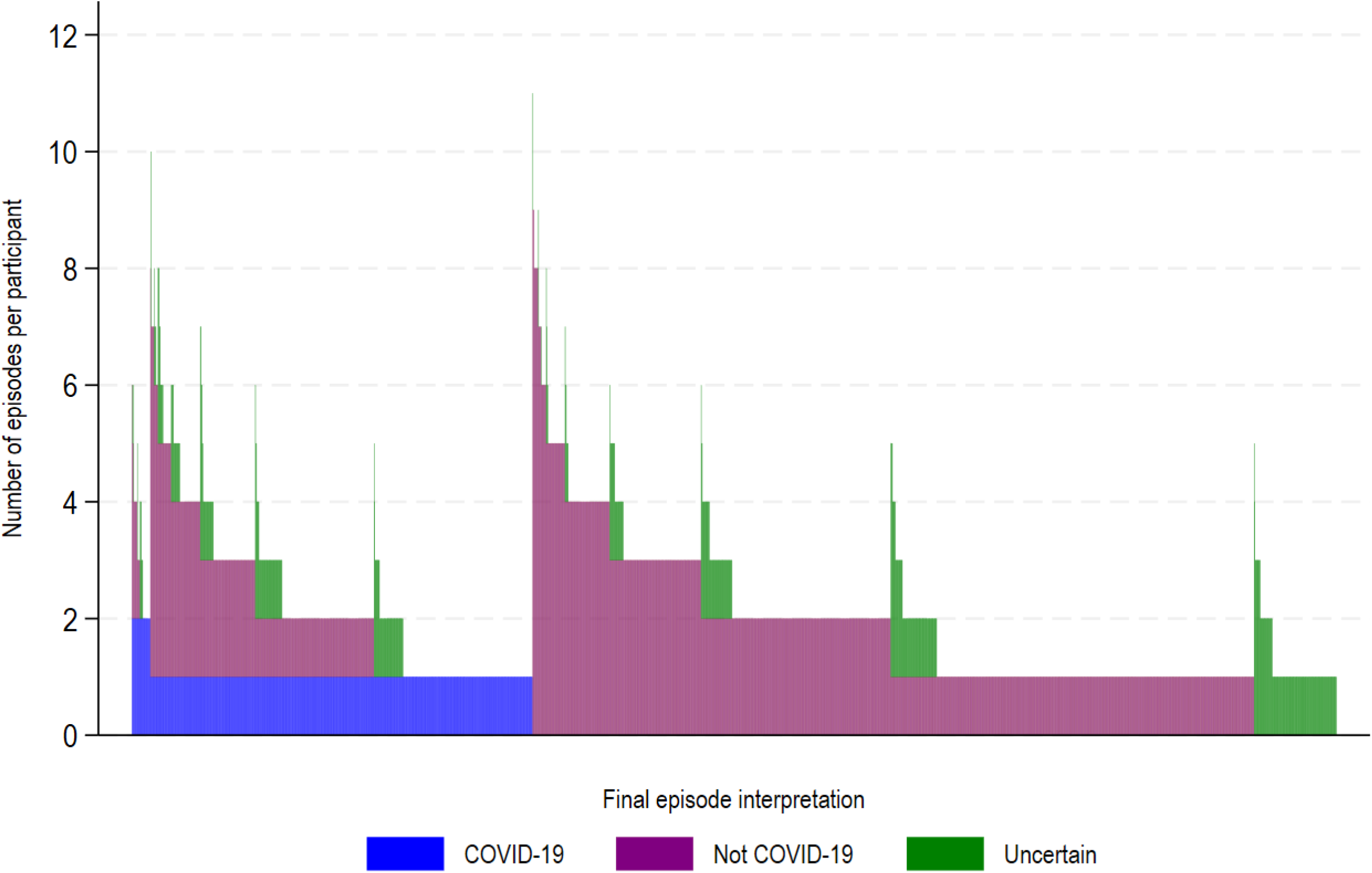
Overall episode interpretation

**Figure 5.**
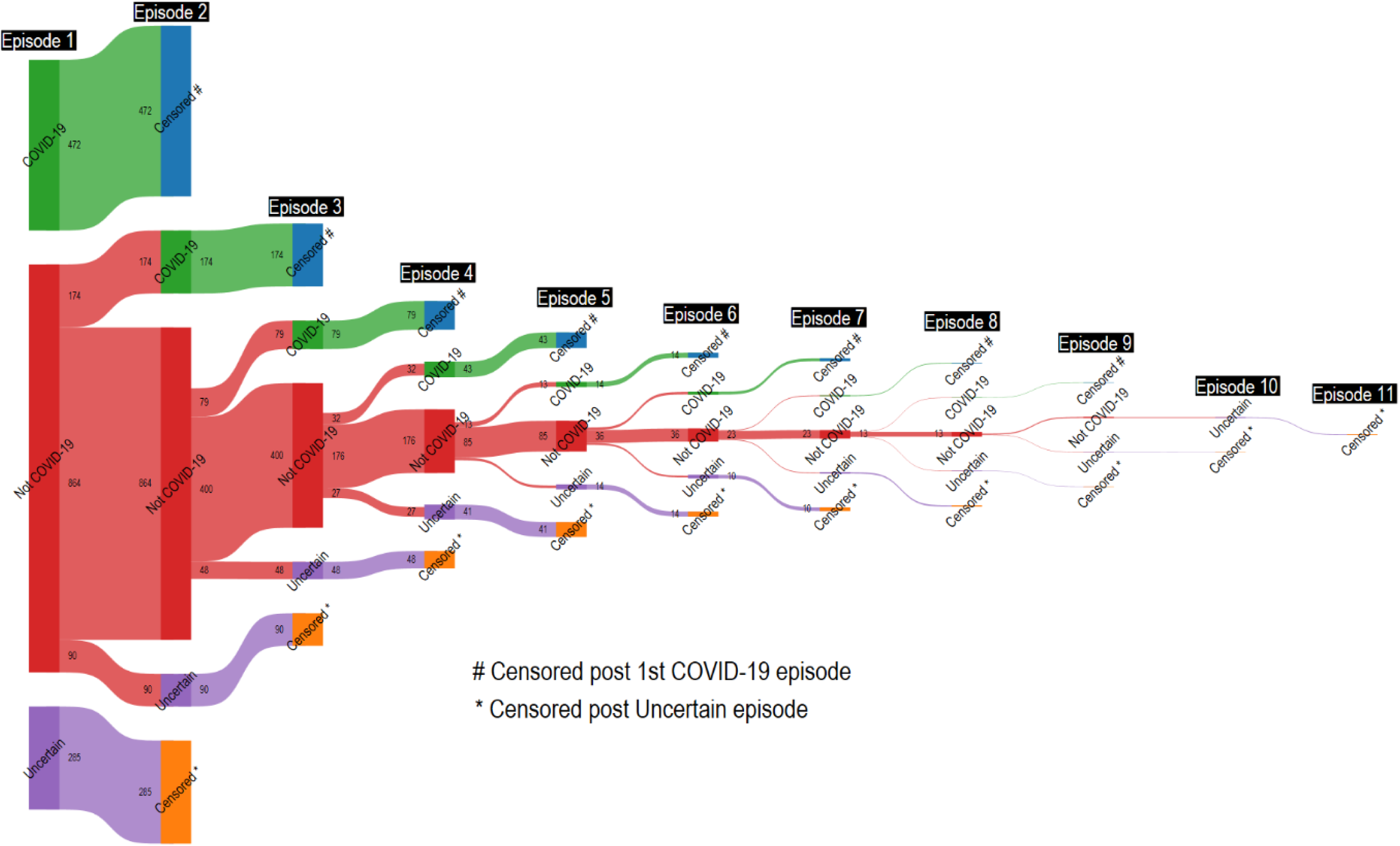
Sequential episode interpretation: COVID-19, Not COVID-19 and Uncertain outcome episodes per participant

## Discussion

The algorithms developed for the BRACE trial provided a novel and robust method for the classification of illness episodes and identification of COVID-19 cases. Unlike other trials that relied exclusively on PCR/RAT [9–11] or serological testing,[12] often classifying cases individually,[13, 14] our three-step algorithm combined these test results with real-world considerations. These included timing of testing relative to symptom onset, variations in the sensitivity and specificity of PCR/RAT and serology, together with the effects of COVID-19 vaccination on serological results.[15] Critically, we accounted for the lower reliability of a negative RAT result due to its lower sensitivity and the impact of *CoronaVac*-induced anti-NCP antibodies.[8]

Post-hoc analysis showed that less than 10% of participants with RAT-negative, PCR-missing episodes seroconverted and were classified as COVID-19, while nearly 15% of participants with RAT-negative, PCR-missing episodes were excluded from primary analysis due to missing serological results.

Discordance between PCR/RAT and serology test results was observed in a small proportion (11%) of episodes. Possible explanations for the 38 (5%) PCR/RAT-positive episodes without seroconversion include mild infections that did not elicit a systemic response, early waning of immunity, limitations in the sensitivity of serology, or false positive PCR/RAT. Possible explanations for the 42 (6%) PCR- negative episodes with seroconversion include inadequate swabbing, technical issues, participant reporting errors, and undetected asymptomatic infections during the interval between blood samples.

Serological testing proved especially valuable when PCR/RAT results were unavailable. Negative serology reclassified 744 episodes with missing PCR/RAT results as Not COVID-19 by demonstrating an absence of seroconversion. Additionally, serology alone identified 11% (98/890) of COVID-19 episodes in which the PCR/RAT result was negative or missing. The algorithm was particularly beneficial in mitigating the impact of incomplete data by reducing the proportion of episodes for which classification would not have been possible (and therefore classified as Uncertain) by more than half, from 30% (1628/5512) to 14% (770/5512).

The addition of serology to PCR/RAT testing involves considerable cost and resources, including staff, consumables, shipment, laboratory assays and data management. However, the benefits included in-person follow-ups that encouraged trial retention, enhanced data accuracy, clarified discordances and the facilitation of additional research opportunities, such as additional immunological studies.[16–21]

Our analyses have some limitations. First, our definition of symptomatic COVID-19 relied on the original case criteria, excluding non-febrile episodes without respiratory symptoms. Second, the timing and type of PCR/RAT testing were not standardised and varied depending on individual settings. Third, serological testing was done quarterly rather than at a consistent interval following illness episodes, which may have affected the detection of seroconversion. Finally, a proportion of illness episodes had incomplete data. However, it is among the COVID-19-related trials with the most complete data available.

Our three-component algorithm developed in the BRACE trial has potential for adaption to diverse epidemiological scenarios beyond COVID-19. Infectious diseases such as influenza or respiratory syncytial virus also require accurate longitudinal monitoring to track transmission, assess vaccine efficacy, and evaluate intervention and treatment outcomes.

In summary, despite robust data collection efforts, COVID-19 trials faced practical challenges, particularly during the early pandemic. Testing availability and participant adherence contributed to the 14% of episodes still classified as missing. Adding serological testing demanded substantial time, financial resources, and participant effort but effectively halved the number of unclassified episodes. Our study underscores the important role of algorithms in achieving comprehensive classification, particularly in complex real-world study settings.

## Supporting information

Supplementary Table_Figures_Consortium

## Data Availability

Deidentified participant data and data dictionary are available to others on request and on completion of a signed data access agreement. Requests can be made in writing to.

## Authors’ Contributions

EM, NC and NM were responsible for the original draft of this paper, with EM responsible for Visualization and analysis. All authors contributed to administration and/or investigation in the BRACE trial, and all were involved in the review and editing of this manuscript. NC, NM, LP and KG were responsible for Conceptualization, Funding acquisition and Supervision. NC, NM, LP and EM were responsible for Methodology. All authors had final responsibility for the decision to submit for publication. NC and NM attest that all listed authors meet authorship criteria and that no others meeting the criteria have been omitted.

## Ethics Committee approval

The study was approved by the Royal Children’s Hospital Melbourne Human Research Ethics Committee (No.62586); the protocol was approved by the ethics committee at each site and all participants provided informed consent. The trial was overseen by a steering committee and a data safety and monitoring board.

## Acknowledgements

We thank the trial participants and site personnel who were involved in assisting the trial (detailed in the Supplementary File). AJ Vaccines (Copenhagen) for facilitating BCG vaccine supplies; Devon Freewheelers (UK) for UK transport; Catalent (UK) for drug management; Fiocruz Clinical Research Platform for coordination and monitoring of sites in Brazil; the Pharmacy, Pathology and Immunisation Service teams from The Royal Children’s Hospital Melbourne, and the Orygen Group for their support.

