## Supplementary Table_Figures_Consortium for "Optimising COVID-19 Episode Identification Using Serology and PCR/Rapid Antigen Testing: Insights from the BRACE Trial"

**Supplementary Table 1.**

| | Seroconverting negative PCR episodes Median (IQR) | Seroconverting positive PCR/RAT episodes Median (IQR) | Wilcoxon rank-sum test Z (Pr) |
|---|---|---|---|
| Post-episode NCP titres | 62.2 (15.9–122.9) | 41.3 (14.3-101.4) | 1.10 (0.27) |
| Time (days) between episode onset and post-episode serological testing | 76.5 (33-102) | 61.0 (37-82) | 1.53 (0.13) |

**Supplementary Figure 1. Serology algorithm**

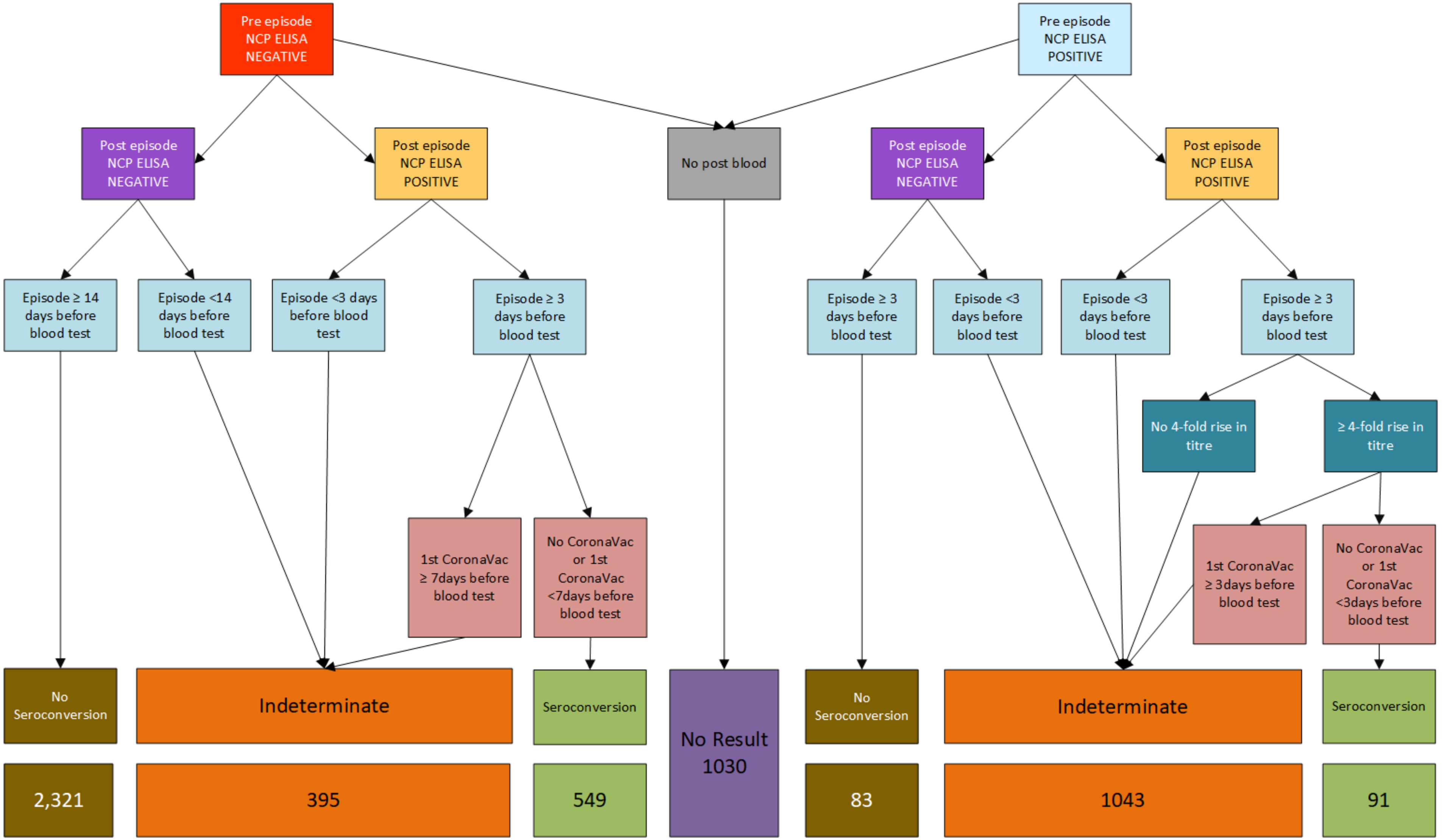

**Supplementary Figure 2. PCR/RAT Interpretation**

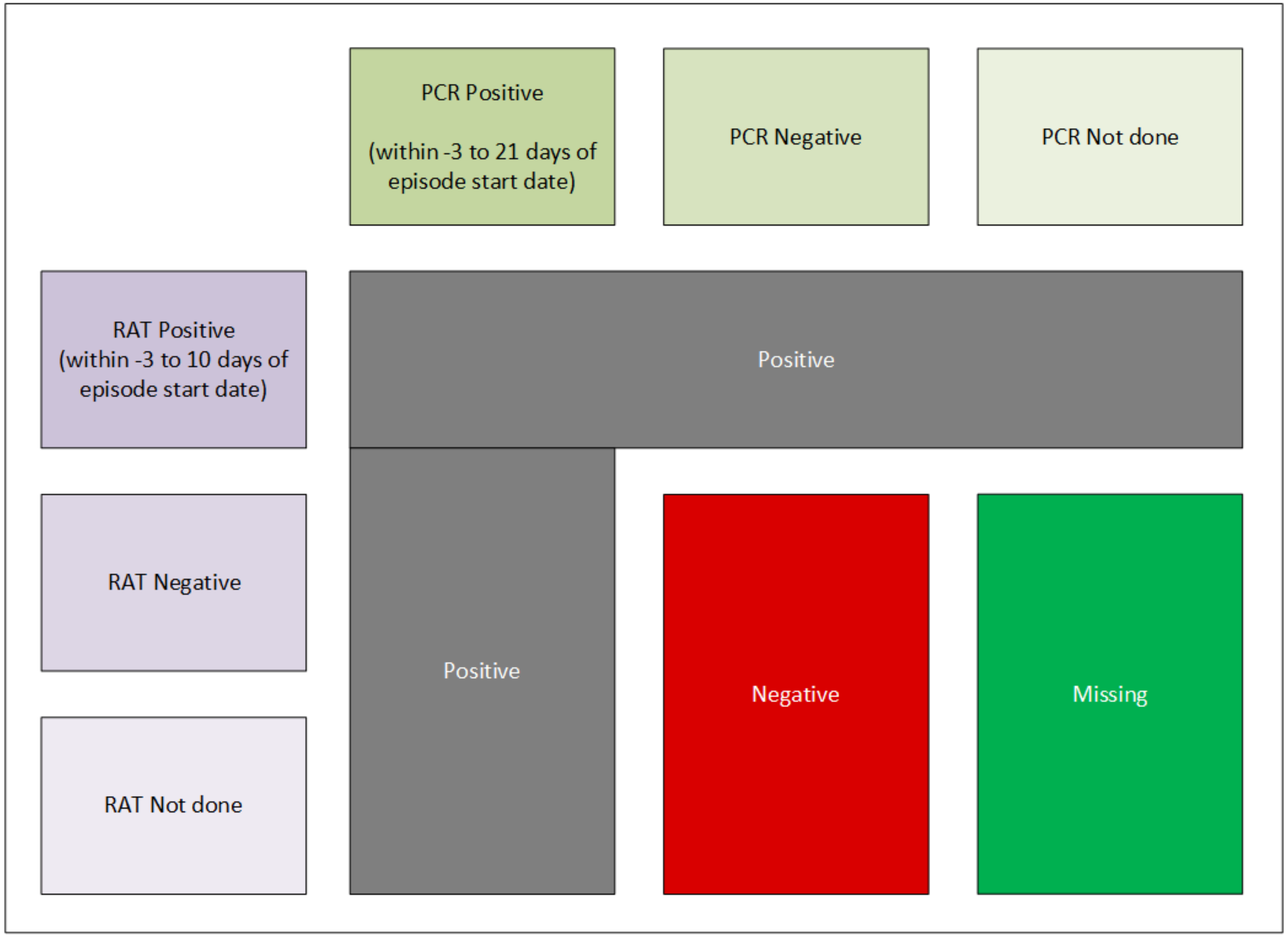

**Supplementary Figure 3. Episode interpretation: pre and post episode NCP results to serology to PCR/RAT to outcome**

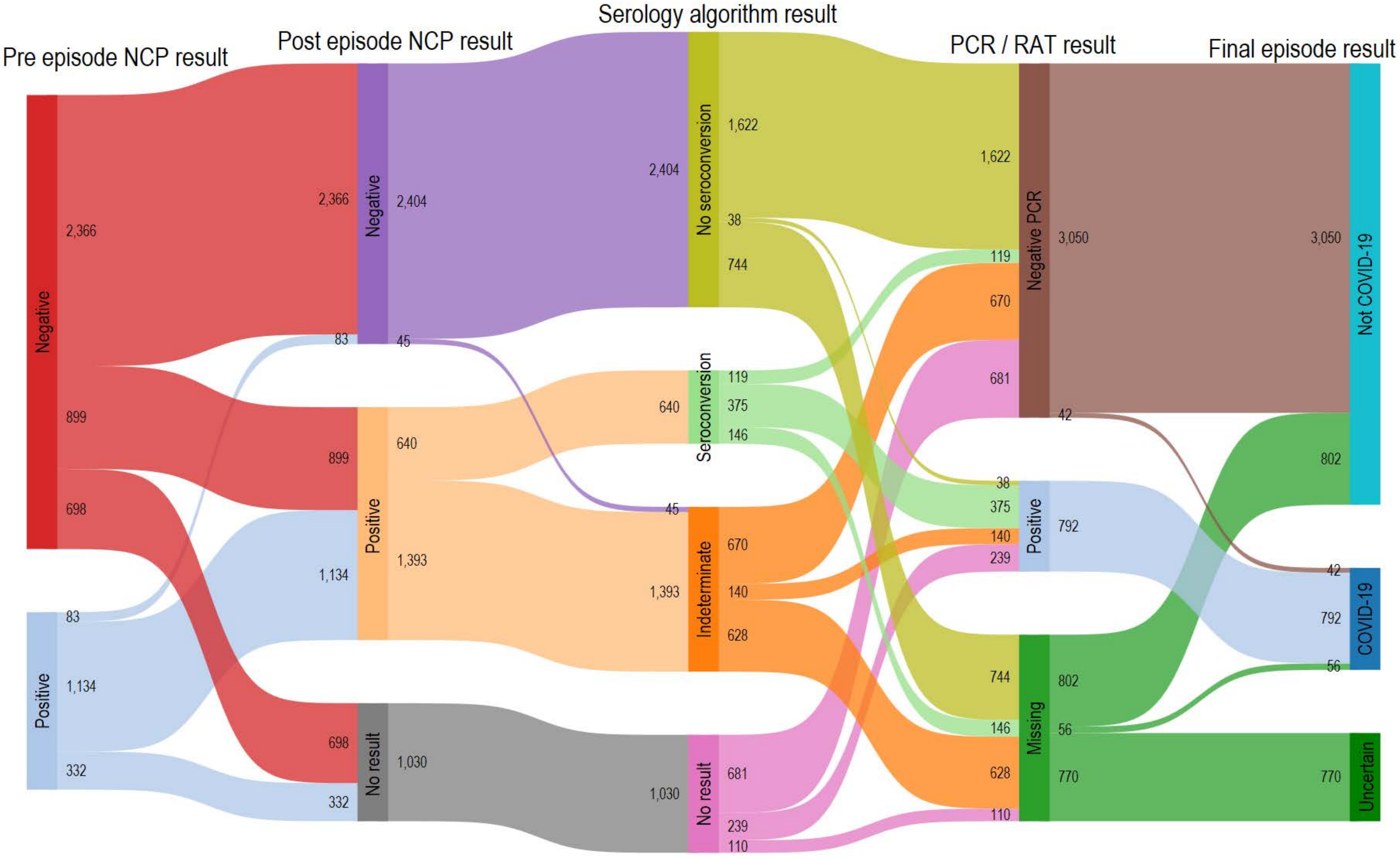

**Supplementary Figure 4. Serology and PCR/RAT test concordance and discordance for COVID-19 episodes**

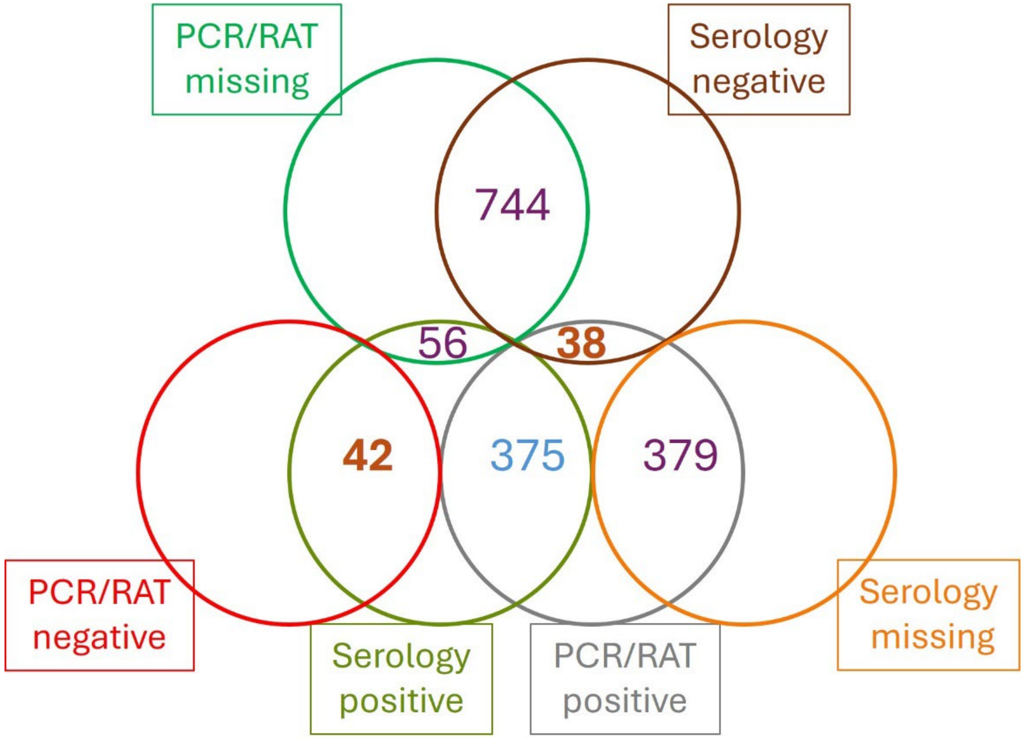

**Supplementary Figure 5. Post-episode NCP titre by days from pre-episode post-episode blood draws for seroconverted PCR negative and PCR/RAT positive episodes**

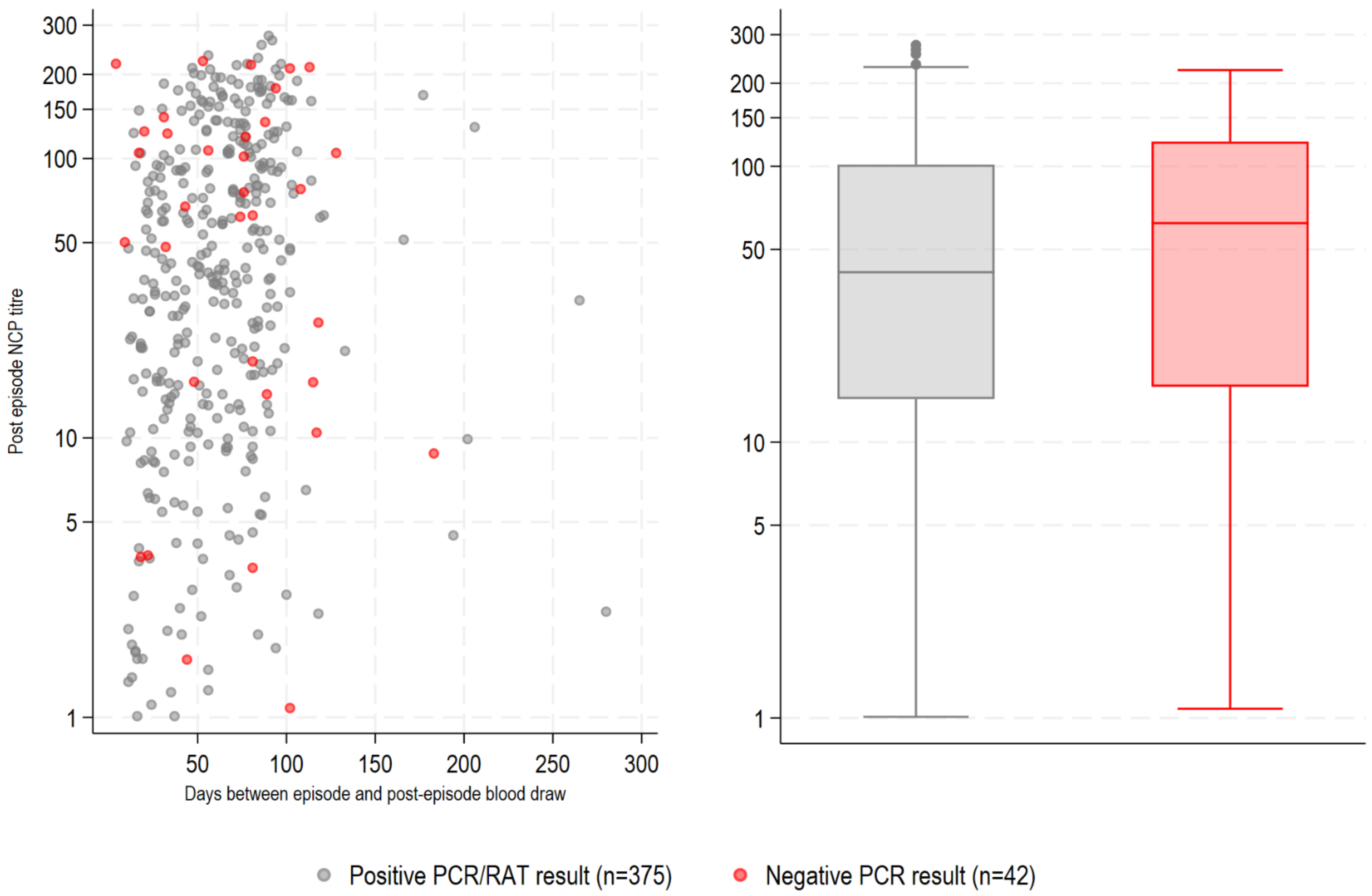

**Supplementary Figure 6. Pre and post NCP titre serology results for episodes with an additional negative rapid antigen test by PCR/RAT and episode result**

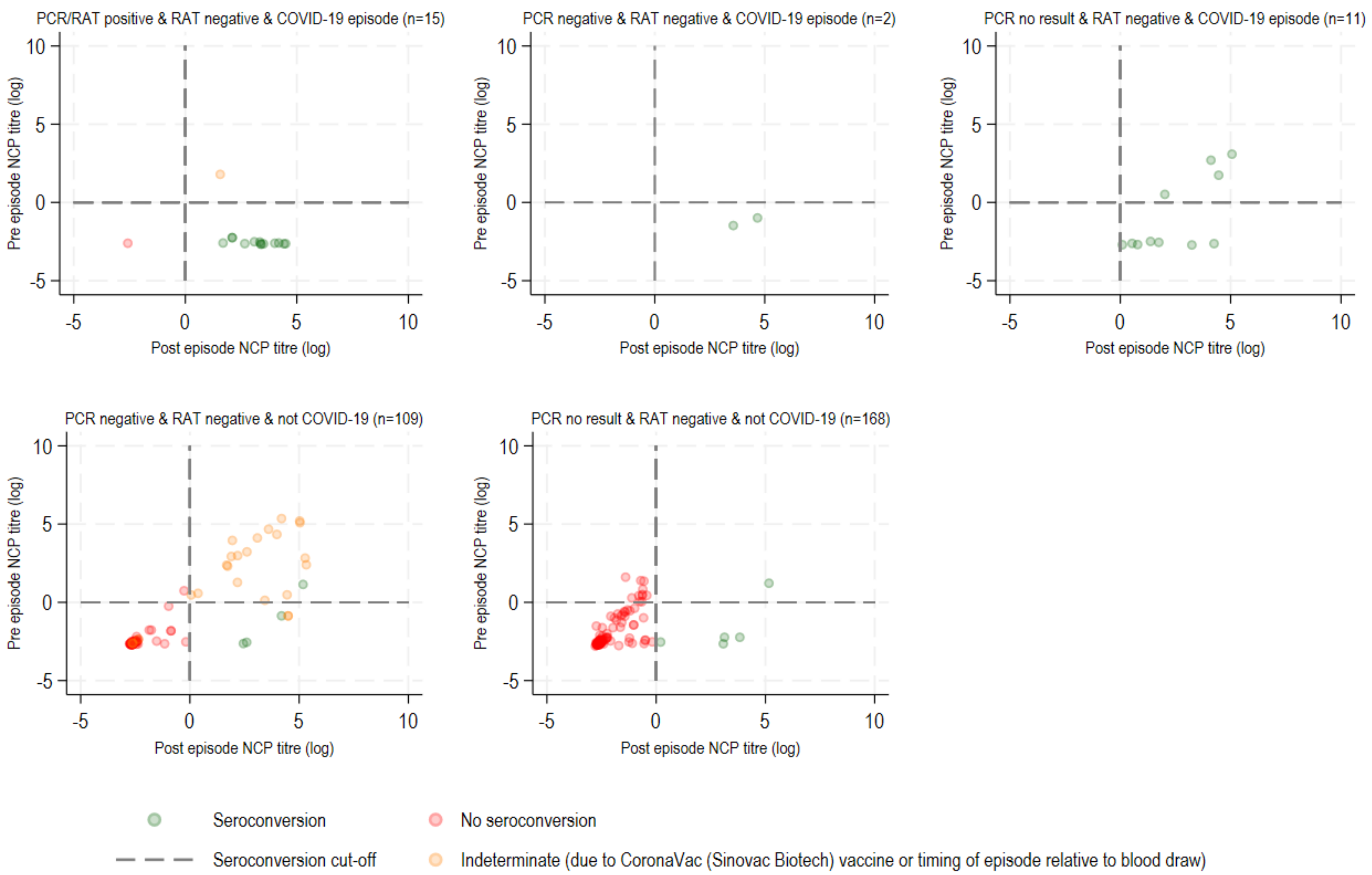

BRACE Trial Consortium Group

### Australia (Victoria)

MCRI Central Team: **Prof Nigel Curtis**, Prof Andrew Davidson, Kaya Gardiner, A/Prof Amanda Gwee, Tenaya Jamieson, Dr Nicole Messina, Thilanka Morawakage, Dr Susan Perlen, A/Prof Kirsten Perrett, Dr Laure Pittet, Amber Sastry, Jia Wei Teo;

Biostatisticians: **Francesca Orsini**, Prof Katherine Lee, Dr Cecilia Moore, Suzanna Vidmar;

Data Team: **Dr Laure Pittet**, Rashida Ali, Ross Dunn, Peta Edler, Grace Gell, Casey Goodall, Richard Hall, Ann Krastev, Dr Nathan La, Dr Ellie McDonald, Nick McPhate, Thao Nguyen, Jack Ren, Luke Stevens;

Laboratory Core Team: **Dr Nicole Messina**, Ahmed Alamrousi, Rhian Bonnici, Dr Thanh Dang, Susie Germano, Jenny Hua, Rebecca McElroy, Monica Razmovska, Scott Reddiex, Xiaofang Wang; Laboratory Scientists: Jeremy Anderson, Kristy Azzopardi, Vicki Bennett- Wood, Anna Czajko, Nadia Mazarakis, Conor McCafferty, Frances Oppedisano, Belinda Ortika, Casey Pell, Leena Spry, Ryan Toh, Sunitha Velagapudi, Amanda Vlahos, Ashleigh Wee-Hee; Biobanking: **Pedro Ramos**, Karina De La Cruz, Dinusha Gamage, Anushka Karunanayake, Isabella Mezzetti, Dr Benjamin Ong, Ronita Singh, Enoshini Sooriyarachchi;

Serology testing (VIDRL): **Dr Suellen Nicholson**, Natalie Cain, Rianne Brizuela, Han Huang;

Study Visit and Phone Call Team: **Veronica Abruzzo**, Morgan Bealing, Patricia Bimboese, Kirsty Bowes, Emma Burrell, Dr Joyce Chan, Jac Cushnahan, Hannah Elborough, Olivia Elkington, Kieran Fahey, Monique Fernandez, Catherine Flynn, Sarah Fowler, Marie Gentile Andrit, Bojana Gladanac, Catherine Hammond, Norine Ma, Sam Macalister, Emmah Milojevic, Jesutofunmi Mojeed, Jill Nguyen, Liz O'Donnell, Nadia Olivier, Isabelle Ooi, Stephanie Reynolds, Lisa Shen, Barb Sherry, Judith Spotswood, Jamie Wedderburn, Angela Younes;

Pharmacy Team: **Donna Legge,** Jason Bell, Jo Cheah, Annie Cobbledick, Kee Lim;

Immunisation Team: **Sonja Elia**, Lynne Addlem, Anna Bourke, Clare Brophy, Nadine Henare, Narelle Jenkins, Francesca Machingaifa, Skye Miller, Kirsten Mitchell, Sigrid Pitkin, Kate Wall;

Safety and Quality Monitoring Team: **Dr Paola Villanueva**, A/Prof Nigel Crawford, Dr Laure Pittet, Dr Wendy Norton;

Epworth Healthcare: **Dr Niki Tan**, Thilakavathi Chengodu, Diane Dawson, Victoria Gordon;

Monash Health: **Tony Korman**, Jess O'Bryan, Veronica Abruzzo;

MCRI Start-up Support Team: Sophie Agius, Dr Samantha Bannister, Jess Bucholc, Alison Burns, Beatriz Camesella, Prof John Carlin, Marianna Ciaverella, Maxwell Curtis, Stephanie Firth, Dr Christina Guo, Matthew Hannan, Erin Hill, Sri Joshi, Katherine Lieschke, Megan Mathers, Sasha Odoi, Ashleigh Rak, Dr Chris Richards, Leah Steve, Carolyn Stewart, Dr Eva Sudbury, Helen Thomson, Emma Watts, Fiona Williams, Angela Young;

Legal: **Penny Glenn**, Andrew Kaynes, Amandine Philippart De Floy App Development: Sandy Buchanan, Thijs Sondag, Ivy Xie;

Media and Communications: **Harriet Edmund**, Bridie Byrne, Tom Keeble, Belle Ngien, Fran Noonan, Michelle Wearing-Smith;

Orygen Volunteers: Alison Clarke, Pemma Davies, Oliver Eastwood, Alric Ellinghaus, Rachid Ghieh, Zahra Hilton, Emma Jennings, Athina Kakkos, Iris Liang, Katie Nicol, Sally O'Callaghan, Helen Osman, Gowri Rajaram, Sophia Ratcliffe, Victoria Rayner, Ashleigh Salmon, Angela Scheppokat, Aimee Stevens, Rebekah Street, Nicholas Toogood.

### Australia (New South Wales)

Westmead Children's Hospital: **A/Prof Nicholas Wood**, Twinkle Bahaduri, Therese Baulman, Jennifer Byrne, Candace Carter, Mary Corbett, Aiken Dao, Maria Desylva, Dr Andrew Dunn, Evangeline Gardiner, Rosemary Joyce, Dr Rama Kandasamy, Prof Craig Munns, Lisa Pelayo, Dr Ketaki Sharma, Katrina Sterling, Caitlin Uren; Westmead Hospital: Clinton Colaco, A/Prof Mark Douglas, Kate Hamilton; Sydney Children's Hospital: Dr Adam Bartlett, Dr Brendan McMullan, Dr Pamela Palasanthiran, Dr Phoebe Williams; Prince of Wales Hospital: Dr Justin Beardsley, Nikki Bergant, Renier Lagunday, Dr Kristen Overton, Prof Jeffrey Post; St Vincent's Hospital Sydney: Dr Yasmeen Al-Hindawi, Sarah Barney, A/Prof Anthony Byrne, Lee Mead, Marshall Plit.

### Australia (South Australia)

SAHMRI: **Prof. David Lynn**, Saoirse Benson, Dr Stephen Blake, Rochelle Botten, Tee Yee Chern, Georgina Eden, Liddy Griffith, Jane James, Dr Miriam Lynn, Angela Markow, Domenic Sacca, Dr Natalie Stevens, Prof. Steve Wesselingh; Royal Adelaide Hospital: Catriona Doran, Dr Simone Barry, Dr Alice Sawka; Women's and Children's Hospital: Dr Sue Evans, Louise Goodchild, Christine Heath, Meredith Krieg, Prof. Helen Marshall, Mark McMillan, Mary Walker.

### Australia (Western Australia)

Perth Children's Hospital/Telethon Kids Institute: **Prof Peter Richmond**, Nelly Amenyogbe, Christina Anthony, Annabelle Arnold, Beth Arrowsmith, Rym Ben-Othman, Sharon Clark, Jemma Dunnill, Nat Eiffler, Krist Ewe, Carolyn Finucane, Lorraine Flynn, Camille Gibson, Lucy Hartnell, Elysia Hollams, Heidi Hutton, Lance Jarvis, Jane Jones, Jan Jones, Karen Jones, Jennifer Kent, Prof Tobias Kollmann, Debbie Lalich, Wenna Lee, Rachel Lim, Sonia McAlister, Fiona McDonald, Andrea Meehan, Asma Minhaj, Lisa Montgomery, Melissa O'Donnell, Jaslyn Ong, Joanne Ong, Kimberley Parkin, Glady Perez, Catherine Power, Shadie Rezazadeh, Holly Richmond, Sally Rogers, Nikki Schultz, Margaret Shave, Patrycja Skut, Lisa Stiglmayer, Alexandra Truelove, Dr Ushma Wadia, Rachael Wallace, Justin Waring; Fiona Stanley Hospital: Michelle England, Erin Latkovic, A/Prof Laurens Manning; Sir Charles Gardiner: Dr Susan Herrmann, Prof Michaela Lucas.

### Brazil (Manaus)

Manaus: **Dr Marcus Lacerda**, Paulo Henrique Andrade, Fabiane Bianca Barbosa, Dayanne Barros, Larissa Brasil, Ana Greyce Capella, Ramon Castro, Erlane Costa, Dilcimar de Souza, Maianne Dias, José Dias, Klenilson Ferreira, Paula Figueiredo, Thamires Freitas, Ana Carolina Furtado, Larissa Gama, Vanessa Godinho, Cintia Gouy, Daniele Hinojosa, Dr Bruno Jardim, Dr Tyane Jardim, Joel Junior, Augustto Lima, Bernardo Maia, Adriana Marins, Kelry Mazurega, Tercilene Medeiros, Rosangela Melo, Marinete Moraes, Elizandra Nascimento, Juliana Neves, Maria Gabriela Oliveira, Thais Oliveira, Ingrid Oliveira, Arthur Otsuka, Rayssa Paes, Handerson Pereira, Gabrielle Pereira, Christiane Prado, Evelyn Queiroz, Laleyska Rodrigues, Bebeto Rodrigues, Dr Vanderson Sampaio, Anna Gabriela Santos, Daniel Santos, Tilza Santos, Evelyn Santos, Ariandra Sartim, Ana Beatriz Silva, Juliana Silva, Emanuelle Silva, Mariana Simão, Caroline Soares, Antonny Sousa, Alexandre Trindade, Dr Fernando Val, Adria Vasconcelos, Heline Vasconcelos.

### Brazil (Mato Grosso do Sul)

Mato Grosso do Sul: **Prof Julio Croda**, Carolinne Abreu, Katya Martinez Almeida, Camila Bitencourt de Andrade, Jhenyfer Thalyta Campos Angelo, Ghislaine Gonçalvez de Araújo Arcanjo, Bianca Maria Silva Menezes Arruda, Wellyngthon Espindola Ayala, Adelita Agripina Refosco Barbosa, Felipe Zampieri Vieira Batista, Fabiani de Morais Batista, Miriam de Jesus Costa, Dr Mariana Garcia Croda, Lais Alves da Cruz, Roberta Carolina Pereira Diogo, Rodrigo Cezar Dutra Escobar, Iara Rodrigues Fernandes, Leticia Ramires Figueiredo, Leandro Galdino Cavalcanti Gonçalves, Sarita Lahdo, Joyce dos Santos Lencina, Guilherme Teodoro de Lima, Larissa Santos Matos, Bruna Tayara Leopoldina Meireles, Debora Quadros Moreira, Lilian Batista Silva Muranaka, Adriely de Oliveira, Karla Regina Warszawski de Oliveira, Matheus Vieira de Oliveira, Prof Roberto Dias de Oliveira, Andrea Antonia Souza de Almeida dos Reis Pereira, Marco Puga, Caroliny Veron Ramos, Thaynara Haynara Souza da Rosa, Karla Lopes dos Santos, Claudinalva Ribeiro dos Santos, Dyenyffer Stéffany Leopoldina dos Santos, Karina Marques Santos, Paulo César Pereira da Silva, Paulo Victor Rocha da Silva, Débora dos Santos Silva, Patricia Vieira da Silva, Bruno Freitas da Rosa Soares, Mariana Gazzoni Sperotto, Mariana Mayumi Tadokoro, Daniel Tsuha, Hugo Miguel Ramos Vieira.

### Brazil (Rio de Janeiro)

Rio de Janeiro: **Prof Margareth Maria Pretti Dalcolmo**, Cíntia Maria Lopes Alves da Paixão, Gabriela Corrêa E Castro, Simone Silva Collopy, Renato da Costa Silva, Samyra Almeida da Silveira, Alda Maria Da-Cruz, Alessandra Maria da Silva Passos de Carvalho, Rita de Cássia Batista, Maria Luciana Silva De Freitas, Aline Gerhardt de Oliveira Ferreira, Ana Paula Conceição de Souza, Paola Cerbino Doblas, Ayla Alcoforado da Silva dos Santos, Vanessa Cristine de Moraes dos Santos, Glauce Dos Santos, Dayane Alves dos Santos Gomes, Anderson Lage Fortunato, Adriano Gomes-Silva, Monique Pinto Gonçalves, Paulo Leandro Garcia Meireles Junior, Estela Martins da Costa Carvalho, Fernando do Couto Motta, Ligia Maria Olivo de Mendonça, Girlene dos Santos Pandine, Rosa Maria Plácido Pereira, Ivan Ramos Maia, Jorge Luiz da Rocha, João Victor Paiva Romano, Erica Fernandes da Silva, Marilda Agudo Mendonça Teixeira de Siqueira, Ágatha Cristinne Prudêncio Soares.

### The Netherlands

UMC Utrecht: **Prof Marc Bonten,** Sandra Franch Arroyo, A/Prof Cristina Prat Aymerich, Henny Ophorst-den Besten, Anna Boon, Karin M Brakke, Axel Janssen, Marijke A.H. Koopmans, Toos Lemmens, Titia Leurink, Engelien Septer-Bijleveld, Kimberly Stadhouders, Dr Darren Troeman, Marije van der Waal, Marjoleine van Opdorp, Nicolette van Sluis, Beatrijs Wolters; Amphia Hospital: Prof Jan Kluytmans, Jannie Romme, Dr Wouter van den Bijllaardt, Linda van Mook, Dr M.M.L (Miranda) van Rijen; Rijnstate Hospital: P. M. G. Filius, Jet Gisolf, Frances Greven, Danique Huijbens, Dr Robert Jan Hassing, R. C. Pon, Lieke Preijers, J. H. van Leusen, Harald Verheij; Noord West Ziekenhuis: Dr Wim Boersma, Evelien Brans, Paul Kloeg, Kitty Molenaar-Groot, Nhat Khanh Nguyen, Dr Nienke Paternotte, Anke Rol, Lida Stooper; Radboud UMC: Helga Dijkstra, Esther Eggenhuizen, Lucas Huijs, Dr Simone Moorlag, Prof Mihai Netea, Eva Pranger, Dr Esther Taks, Dr Jaap ten Oever, Rob ter Heine; St Antonius Hospital: Kitty Blauwendraat, Dr Bob Meek, Isil Erkaya, Houda Harbech, Dr Nienke Roescher, Rifka Peeters, Menno te Riele, Carmen Zhou.

### Spain

Mutua Terrassa University Hospital: Dr Esther Calbo, Cristina Badia Marti, Emma Triviño Palomares, Tomás Perez Porcuna; University Hospital Germans Trias I Pujol: Anabel Barriocanal, Ana Maria Barriocanal, Irma Casas, Jose Dominguez, Maria Esteve, Alicia Lacoma, Irene Latorre, Gemma Molina, Barbara Molina, Dr Antoni Rosell, Sandra Vidal; Hospital Virgen Macarena: Lydia Barrera, Natalia Bustos, Ines Portillo Calderón, David Gutierrez Campos, Jose Manuel Carretero, Angel Dominguez Castellano, Renato Compagnone, Encarnacion Ramirez de Arellano, Almudena de la Serna, Maria Dolores del Toro Lopez, Marie-Alix Clement Espindola, Ana Belen Martin Gutierrez, Alvaro Pascual Hernandez, Virginia Palomo Jiménez, Elisa Moreno, Nicolas Navarrete, Teresa Rodriguez Paño, Prof Jesús Rodríguez-Baño, Enriqueta Tristán, Maria Jose Rios Villegas; University Hospital Cruces: Atsegiñe Canga Garces, Erika Castro Amo, Raquel Coya Guerrero, Dr.Josune Goikoetxea, Leticia Jorge, Cristina Perez; Marqués de Valdecilla University Hospital: Dr María Carmen Fariñas Álvarez, Manuel Gutierrez Cuadra, Dr Francisco Arnaiz de las Revillas Almajano, Pilar Bohedo Garcia, Dr Teresa Giménez Poderos, Claudia González Rico, Blanca Sanchez, Olga Valero, Noelia Vega.

### United Kingdom

University of Exeter/Exeter Clinical Trials Unit: **Prof John Campbell,** Anna Barnes, Dr Helen Catterick, Tim Cranston, Phoebe Dawe, Emily Fletcher, Liam Fouracre, Dr Alison Gifford, Neil Gow, John Kirkwood, Dr Christopher Martin, Dr Amy McAndrew, Marcus Mitchell, Georgina Newman, Dr Abby O'Connell, Jakob Onysk, Lynne Quinn, Dr Shelley Rhodes, Samuel Stone, Dr Lorrie Symons, Harry Tripp, Prof Adilia Warris, Darcy Watkins, Bethany Whale; St Leonard's Practice: Dr Alex Harding, Gemma Lockhart, Dr Kate Sidaway-Lee; Ide Lane Surgery: Dr John Campbell, Dr Sam Hilton, Sarah Manton, Dr Daniel Webber-Rookes, Rachel Winder; Travel Clinic: James Moore; Royal Devon and Exeter NHS Foundation Trust: Freya Bateman, Dr Michael Gibbons, Dr Bridget Knight, Julie Moss, Dr Sarah Statton, Josephine Studham; Teign Estuary Medical Group/Glendevon Medical Practice: Lydia Hall, Will Moyle, Dr Tamsin Venton.
